# Influencers, not just adverts: social media influencer exposure and tobacco use among urban youth in Kampala and Nairobi — a comparative mixed-methods study

**DOI:** 10.64898/2026.06.06.26355037

**Authors:** Jeya Sushimitha Jawahar Kanth, Thuparambil M. Ravi Anish, Benjamin Odhiambo, Kiragga Denis Lwembawo, Segawa Micheal, Jim Arinaitwe, Lydia Nakiyingi

**Author notes:** **Corresponding author:** Jeya Sushimitha Jawahar Kanth, Centre for Tobacco Control in Africa / Makerere University, Kampala, Uganda. **Co-author emails:**.

## Abstract

**Background:** Tobacco control treaties were written for billboards and television, not for the people now selling lifestyles to young Africans. As mobile internet saturates East African cities, social media influencers have become an unmeasured channel, especially when it comes to tobacco promotion. We assessed the prevalence of tobacco use, its association with influencer exposure, and how urban youth interpret that exposure in two capitals with different tobacco laws.

**Methods:** We conducted a comparative mixed-methods study among youth aged 18–29 years in Kampala, Uganda, and Nairobi, Kenya (January–August 2025), combining (i) a cross-sectional survey using systematic sampling at youth-dense venues (n=772), (ii) four online focus group discussions (FGDs; n=40), and (iii) content analysis of 30 tobacco-related posts from high-reach influencers (≥50,000 followers). We used chi-square tests and multivariable logistic regression, thematic analysis (Braun and Clarke), and descriptive engagement metrics.

**Results:** Ever tobacco use among urban youth in East Africa was 29.3% (226/772), similar in Kampala (30.7%) and Nairobi (28.0%; p=0.409). After adjustment, exposure to influencers promoting tobacco independently predicted ever use (adjusted odds ratio [aOR] 1.90, 95% confidence interval [CI] 1.29– 2.82; p=0.001), alongside male sex (aOR 2.35) and age 26–29 years (aOR 1.99). Tertiary education (aOR 0.45) and never seeing tobacco content (aOR 0.26) were protective. Posts framed tobacco as aspirational lifestyle; 77% of sampled comments were positive and 47.5% expressed interest in trying the product.

**Conclusions:** Influencer exposure behaved as a modifiable risk factor of a magnitude comparable to established demographic drivers. Tobacco control in the region must move from print-era advertising bans to platform governance, mandatory disclosure of paid promotion, and youth-led counter-marketing.

**What this paper adds:**

- **What is already known:** High-income studies link social media tobacco content to youth use, but African evidence is largely anecdotal and rarely distinguishes generic exposure from influencer-mediated promotion.
- **What this study adds:** In two East African capitals, influencer exposure was independently associated with ever tobacco use after adjustment, and triangulated qualitative and content data show how aspirational framing converts reach into intent.
- **Implications:** Existing advertising bans do not reach influencer content; regulation, disclosure rules and credible counter-voices are needed.

## Introduction

The instruments that govern tobacco marketing were built for a world of billboards, point-of-sale displays and television spots. They were not built for a twenty-three-year-old in Kampala watching a favourite influencer exhale flavoured vapour in a rooftop bar, or for the Nairobi student who first sees shisha not in an advertisement but in a friend-of-a-friend’s holiday reel. Yet this is where a growing share of tobacco promotion now happens, and it is precisely the space where most tobacco control law in sub-Saharan Africa falls silent.

The scale of the shift is not in dispute. More than five billion people now use social media, and young adults aged 18–29 years spend an estimated three to four hours each day on platforms such as Instagram, TikTok and YouTube (DataReportal, 2024). In sub-Saharan Africa, mobile connectivity is now near-universal among the young: roughly four in five people aged 15–24 years are internet users (International Telecommunication Union, 2023). Into this environment have stepped social media influencers — people who convert a dedicated online following into commercial persuasion through lifestyle content and product placement. Their power lies less in overt salesmanship than in the perception that they are relatable and trustworthy peers (Abidin, 2016; De Veirman, Cauberghe and Hudders, 2017). Where followers believe an endorsement is authentic rather than paid, its persuasive force increases (Dhanesh and Duthler, 2019) — a detail that matters greatly when the product is tobacco and the disclosure is absent.

Tobacco remains one of the largest preventable causes of death, killing in the order of eight million people each year, with about 80% of the world’s 1.3 billion users living in low- and middle-income countries (World Health Organization, 2023). The WHO Framework Convention on Tobacco Control (FCTC) curtailed conventional advertising, but the digital ecosystem has opened as a largely ungoverned frontier in which tobacco imagery circulates freely (World Health Organization, 2025). This matters most where the market is youngest and the rules are weakest. Sub-Saharan Africa has the world’s lowest smoking prevalence but its fastest-growing number of smokers — projected to rise from 66 million in 2015 to 84 million by 2025 — making the region the probable epicentre of the next tobacco epidemic (Egbe, Bialous and Glantz, 2022). Roughly 65% of Africa’s population is under 30 years (United Nations Population Fund, 2023), and the industry has every commercial reason to reach them where they already are.

The evidence linking social media to youth tobacco use is, by now, robust — but it is overwhelmingly drawn from high-income settings. A systematic review and meta-analysis of 29 studies found that exposure to tobacco content on social media roughly doubled the odds of lifetime tobacco use (odds ratio [OR] 2.18, 95% confidence interval [CI] 1.54–3.08) and of susceptibility among never-users (Donaldson et al., 2022). Comparable associations have been reported for e-cigarette content specifically (Ghahramani, Ratan and Ahmed, 2022). These figures are persuasive, but they are not automatically transferable. African urban contexts differ in regulatory capacity, in the dominance of mobile-first access, in the cultural meaning attached to shisha and flavoured products, and in the degree to which enforcement extends to online space. Uganda’s Tobacco Control Act (2015) and Kenya’s Tobacco Control Act (2007) both prohibit tobacco advertising, yet neither is meaningfully enforced against influencer content, and civil-society reporting suggests active recruitment of online personalities to promote nicotine products (Kenya Tobacco Control Alliance, 2024; Majiwa, 2024).

Two gaps follow. First, almost no African study distinguishes generic social media use from influencer-mediated promotion — a distinction that determines which policy lever applies. Second, the existing literature rarely triangulates what young people are exposed to, how they interpret it, and how that content actually performs. We addressed both by combining a survey, focus groups and content analysis in two capitals with deliberately different policy histories. We aimed to (i) estimate the prevalence of ever tobacco use among urban youth, (ii) test whether exposure to influencers promoting tobacco is independently associated with use after adjustment for established determinants, and (iii) characterise the content, reach and reception of pro-tobacco influencer messaging.

## Methods

### Design and setting

We conducted a comparative, convergent mixed-methods study integrating a cross-sectional survey, focus group discussions (FGDs) and a structured social media content analysis. Kampala and Nairobi were selected for their large, digitally engaged youth populations and for their contrasting tobacco control trajectories, allowing a natural comparison of two regulatory environments facing a shared digital exposure. Reporting follows the Strengthening the Reporting of Observational Studies in Epidemiology (STROBE) and Standards for Reporting Qualitative Research (SRQR) guidelines. Data were collected between January and August 2025.

### Population and sampling

Eligible participants were youth aged 18–29 years resident in Kampala or Nairobi who could provide informed consent; those with cognitive impairment precluding participation were excluded. Using Schlesselman’s formula (95% confidence, 80% power, baseline prevalence 11%, assumed OR 2.0), we required 374 participants per city (total 748). We used systematic sampling at youth-dense venues — university campuses, community centres, youth clubs and public parks — selecting every nth person entering each venue, with n fixed randomly at the start of collection. Eligible individuals gave electronic consent and self-completed a questionnaire on a mobile device, with research assistants available for technical support only.

For the qualitative component, we purposively recruited survey participants who reported active engagement with influencers, conducting four FGDs (two per city, n=40) via Google Meet. Sessions were audio-recorded with consent, facilitated with a semi-structured guide and transcribed verbatim. For content analysis, we purposively assembled 30 tobacco-related posts from high-reach influencers (≥50,000 followers) identified through tobacco-related hashtags (e.g. #Shisha, #Tobacco) on Instagram, TikTok, Twitter (X) and YouTube. The post — not the individual — was the unit of analysis, because engagement, framing and product type attach to posts rather than to people.

### Measures

The primary outcome was ever tobacco use: self-reported use of any tobacco or nicotine product (cigarettes, shisha, e-cigarettes, smokeless tobacco) at least once. Independent variables comprised sociodemographic factors (age, sex, education, city, employment, living arrangement), digital exposure (daily time on social media, frequency of seeing tobacco content, following influencers, seeing influencers promote tobacco) and perception variables (whether influencers make tobacco appealing; self-rated likelihood of trying tobacco after such exposure; perceived peer pressure). For each post, we coded influencer reach, content type, product featured, message theme and tone, appeal strategy and engagement metrics (views, likes, comments, shares); comment sentiment was classified as positive, critical or neutral.

### Analysis

Quantitative data were analysed in R version 4.5.1. Categorical variables were summarised as frequencies and percentages and compared using Pearson’s chi-square tests. Variables associated with the outcome at p<0.20 in univariable analysis entered a multivariable logistic regression model. Multicollinearity was checked using the variance inflation factor (VIF) and model fit assessed using McFadden’s pseudo-R^2^, the Akaike information criterion (AIC) and deviance. Adjusted odds ratios (aOR) with 95% CIs are reported; significance was set at p≤0.05. FGD transcripts were analysed thematically following Braun and Clarke’s (2006) six-step framework, coded inductively and deductively in ATLAS.ti version 25 by two independent coders with discrepancies resolved by consensus. Engagement metrics were summarised as medians and interquartile ranges (IQR), and compared across platforms and content strategies.

### Ethics

Approval was granted by the Makerere University School of Public Health Research Ethics Committee (SPH-2024-302) and the KCA University Research Ethics Committee (KCAU-SERC-EXTER003), with regulatory registration by the Uganda National Council for Science and Technology (SS3850ES) and Kenya’s National Commission for Science, Technology and Innovation (NACOSTI/P/25/4174349). All participants gave electronic informed consent. Data were anonymised and stored securely. The study followed the Declaration of Helsinki.

## Results

### Sample and prevalence

Of 772 participants (375 Kampala; 397 Nairobi), most were aged 22–25 years (54%), male (54%) and tertiary-educated (87%); two-thirds were unemployed (Table 1). Ever tobacco use was 29.3% (226/772) and did not differ between cities (Kampala 30.7% vs Nairobi 28.0%; p=0.409). The near-identical prevalence across two different legal regimes is itself informative: shared digital exposure appears to override divergent national policy on paper. Almost half (47%) spent more than four hours daily on social media, 39% had seen influencers promote tobacco, and 46% agreed influencers make tobacco appealing — yet 52% judged themselves unlikely to try it after such exposure, signalling a reservoir of resistance that prevention can build on.

**Table 1.**
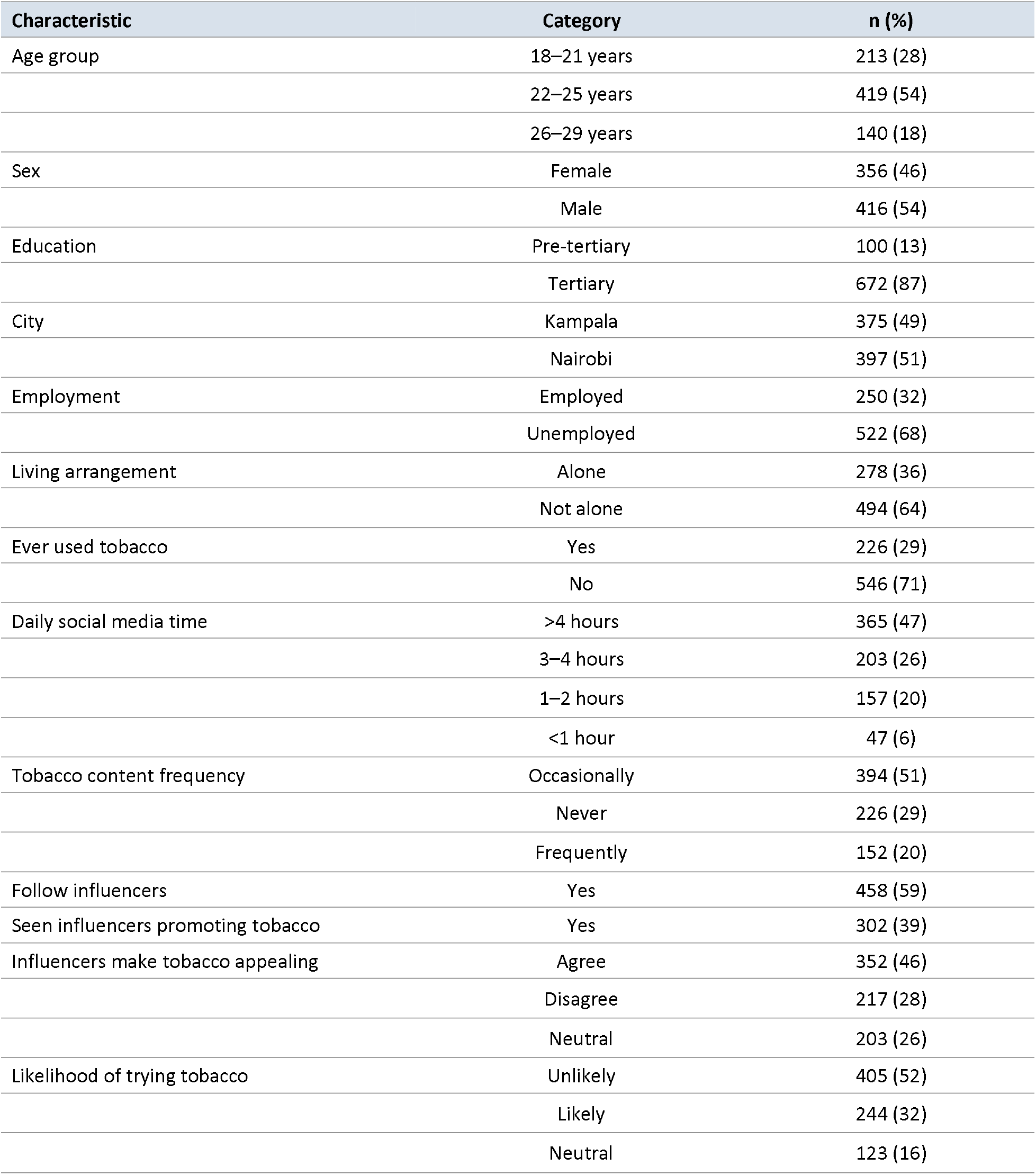
Sociodemographic characteristics and key variables (n=772)

| Characteristic | Category | n (%) |
| --- | --- | --- |
| Age group | 18–21 years | 213 (28) |
|  | 22–25 years | 419 (54) |
|  | 26–29 years | 140 (18) |
| Sex | Female | 356 (46) |
|  | Male | 416 (54) |
| Education | Pre-tertiary | 100 (13) |
|  | Tertiary | 672 (87) |
| City | Kampala | 375 (49) |
|  | Nairobi | 397 (51) |
| Employment | Employed | 250 (32) |
|  | Unemployed | 522 (68) |
| Living arrangement | Alone | 278 (36) |
|  | Not alone | 494 (64) |
| Ever used tobacco | Yes | 226 (29) |
|  | No | 546 (71) |
| Daily social media time | >4 hours | 365 (47) |
|  | 3–4 hours | 203 (26) |
|  | 1–2 hours | 157 (20) |
|  | <1 hour | 47 (6) |
| Tobacco content frequency | Occasionally | 394 (51) |
|  | Never | 226 (29) |
|  | Frequently | 152 (20) |
| Follow influencers | Yes | 458 (59) |
| Seen influencers promoting tobacco | Yes | 302 (39) |
| Influencers make tobacco appealing | Agree | 352 (46) |
|  | Disagree | 217 (28) |
|  | Neutral | 203 (26) |
| Likelihood of trying tobacco | Unlikely | 405 (52) |
|  | Likely | 244 (32) |
|  | Neutral | 123 (16) |

### Independent predictors of tobacco use

The model fitted acceptably (McFadden pseudo-R^2^=0.184; AIC=793.63; deviance=761.63) with no multicollinearity (all VIF<5). Three findings stand out (Table 2). First, exposure to influencers promoting tobacco independently predicted ever use (aOR 1.90, 95% CI 1.29–2.82; p=0.001) after adjustment for demographic and behavioural confounders. The crude association was markedly stronger (crude OR 3.11), and its attenuation under adjustment is reassuring rather than disappointing: it indicates that part of the raw signal reflected confounding, while a substantial, independent association survived. Second, the magnitude of that adjusted effect is comparable to classic determinants — male sex (aOR 2.35, 95% CI 1.62–3.42) and age 26–29 years (aOR 1.99, 95% CI 1.14–3.48) — which reframes influencer exposure not as a peripheral curiosity but as a determinant of similar order. Third, our point estimate sits squarely within the range of the pooled high-income estimate (OR 2.18) reported by Donaldson et al. (2022), suggesting the mechanism travels across very different markets.

**Table 2.** Multivariable predictors of ever tobacco use among urban youth (n=772)

| Variable | Category | aOR | 95% CI | p-value |
| --- | --- | --- | --- | --- |
| Age group | 18–21 (Ref) | 1.00 | — | — |
|  | 22–25 | 1.07 | 0.70–1.66 | 0.759 |
|  | 26–29 | 1.99 | 1.14–3.48 | 0.015 |
| Sex | Female (Ref) | 1.00 | — | — |
|  | Male | 2.35 | 1.62–3.42 | <0.001 |
| Education | Pre-tertiary (Ref) | 1.00 | — | — |
|  | Tertiary | 0.45 | 0.27–0.75 | 0.002 |
| City | Kampala (Ref) | 1.00 | — | — |
|  | Nairobi | 0.92 | 0.64–1.32 | 0.652 |
| Employment | Employed (Ref) | 1.00 | — | — |
|  | Unemployed | 0.74 | 0.50–1.10 | 0.132 |
| Living arrangement | Not alone (Ref) | 1.00 | — | — |
|  | Alone | 1.16 | 0.79–1.72 | 0.447 |
| Daily social media time | 1–2 h (Ref) | 1.00 | — | — |
|  | <1 h | 2.39 | 1.04–5.53 | 0.041 |
|  | 3–4 h | 1.13 | 0.72–1.79 | 0.599 |
|  | >4 h | 1.49 | 0.92–2.42 | 0.108 |
| Tobacco content frequency | Frequently (Ref) | 1.00 | — | — |
|  | Never | 0.26 | 0.14–0.47 | <0.001 |
|  | Occasionally | 0.64 | 0.42–0.98 | 0.042 |
| Follow influencers | No (Ref) | 1.00 | — | — |
|  | Yes | 0.77 | 0.51–1.15 | 0.201 |
| Influencers promote tobacco | No (Ref) | 1.00 | — | — |
|  | Yes | 1.90 | 1.29–2.82 | 0.001 |
| Influencers appealing | Agree (Ref) | 1.00 | — | — |
|  | Disagree | 1.26 | 0.77–2.06 | 0.364 |
|  | Neutral | 1.28 | 0.79–2.07 | 0.312 |
| Likelihood of trying tobacco | Likely (Ref) | 1.00 | — | — |
|  | Neutral | 0.73 | 0.44–1.21 | 0.221 |
|  | Unlikely | 0.30 | 0.19–0.47 | <0.001 |
| Peer pressure | No (Ref) | 1.00 | — | — |
|  | Yes | 1.44 | 0.97–2.13 | 0.071 |

aOR, adjusted odds ratio; CI, confidence interval; Ref, reference category. Model fit: McFadden pseudo- $R^2=0.184$ ; AIC=793.63; deviance=761.63; all VIF<5.

Protective associations were equally instructive. Tertiary education more than halved the odds of ever use (aOR 0.45, 95% CI 0.27–0.75), and a clear exposure gradient emerged: never seeing tobacco content (aOR 0.26) and occasional exposure (aOR 0.64) were both protective relative to frequent exposure, while reporting oneself unlikely to try tobacco after seeing an influencer use it was strongly protective (aOR 0.30). Read together, these point to two distinct, actionable levers — reducing the volume of exposure and strengthening resistance to it.

One counter-intuitive result deserves honest treatment rather than burial. Spending less than one hour daily on social media was associated with higher odds of ever use than spending one to two hours (aOR 2.39, 95% CI 1.04–5.53). This almost certainly reflects who the light users are — older, employed men whose tobacco use predates and is unrelated to heavy platform engagement — rather than a protective effect of scrolling. It is a useful caution against equating total screen time with risk: the relevant exposure is tobacco-specific content, not time online per se, a distinction the content-frequency gradient confirms.

### How youth read influencer content

Four themes emerged from the FGDs. Pathways to use began early and socially — childhood curiosity and familial normalisation, later reinforced by peers in campus and nightlife settings. Influencers as normalisation agents was the most consistent theme: participants described smoking and especially vaping being folded into aspirational lifestyles rather than openly advertised, so that promotion was felt but rarely named. One Ugandan participant captured the parasocial pull plainly: “When you see your favourite influencer using shisha in a nice place, you want to try it too.” Product-specific risk rationalisation revealed a coherent but dangerous hierarchy of harm — cigarettes seen as “dirty” and addictive, shisha and vapes as cleaner, modern and safer — that maps directly onto industry framing of novel products as harm-reduction. Counter-narratives showed agency: participants proposed enlisting trusted influencers for anti-tobacco messaging, short-form educational content, platform regulation and media literacy, several explicitly wanting it “trendy to be smoke-free.” Cross-country nuance mattered: Kenyan participants foregrounded digital and influencer drivers, whereas Ugandan participants emphasised peer and community contexts — a difference that argues against one-size-fits-all regional campaigns.

### What the content does

The 30 sampled posts were predominantly video (63%) and almost universally framed tobacco within a lifestyle theme (93%); e-cigarettes/vapes were the most featured product (60%). Reach was substantial and skewed (median 220,482 views, IQR 1,845,073), with the highest median viewership on YouTube (1,834,659) and TikTok (803,350) (Table 3). Reception was overwhelmingly favourable: a median 77% of comments were positive and 47.5% expressed interest in trying the featured product, rising to 90% interest on Instagram. Strategy mattered more than frequency. Celebrity endorsement and status symbolism were the most common devices, yet emotional appeal and humour were the most effective, generating far higher median views (4.83 million and 0.31 million respectively). The practical lesson is uncomfortable for conventional health messaging: it is not exposure alone but emotionally resonant, entertaining framing that converts reach into intent — and that is exactly the register counter-marketing must learn to match.

**Table 3.** Reach and reception of pro-tobacco influencer posts, by platform (n=30 posts)

| Metric | Overall | Instagram | TikTok | Twitter (X) | YouTube |
| --- | --- | --- | --- | --- | --- |
| Posts, n | 30 | 6 | 6 | 12 | 6 |
| Median followers | 174,500 | 182,250 | 568,250 | 72,700 | 196,500 |
| Median views | 220,482 | 1,475,000 | 803,350 | 27,650 | 1,834,659 |
| Median likes | 7,239 | 81,950 | 62,456 | 377 | 31,000 |
| Median comments | 103 | 198 | 176 | 7 | 958 |
| Positive comments (%) | 77 | 80 | 80 | 80 | 60 |
| Interest in trying (%) | 47.5 | 90 | 46 | 20 | 45 |

*Engagement values are medians. Video content predominated (63%); lifestyle framing predominated (93%); e-cigarettes/vapes were the most featured product (60%).*

## Discussion

This study provides some of the first African evidence that exposure to influencers promoting tobacco is independently associated with tobacco use among urban youth, and that the association is of a magnitude comparable to sex and age. The convergence of three methods is the central strength: the survey quantifies the association, the FGDs explain the mechanism (aspirational, parasocial normalisation), and the content analysis shows that the material doing the work is highly engaging and warmly received.

Our ever-use prevalence (29.3%) is considerably higher than national adult estimates such as Uganda’s ∼7–10% (Kabwama et al., 2018; Ministry of Health Uganda, 2021). This is expected rather than contradictory: national figures pool rural and urban populations and often measure current rather than ever use, whereas we sampled digitally engaged youth in capital cities, where autonomy, disposable income, retail density and online exposure concentrate. The near-identical prevalence in Kampala and Nairobi, despite different statutes, reinforces a regional reading: a shared digital environment is producing convergent youth behaviour that national legislation, written for offline marketing, does not touch.

The finding that demands action is the product hierarchy. Participants treated shisha and vapes as cleaner and safer than cigarettes — precisely the perception the industry cultivates for novel products and disseminates through covert influencer placement that evades platform advertising bans. African tobacco control frameworks were drafted for combustible cigarettes; they are poorly equipped for flavoured vapes and shisha promoted as lifestyle accessories (Egbe, Bialous and Glantz, 2022). That regulatory lag, colliding with the highest engagement metrics in our sample being vape content, points to an urgent need to update both law and public messaging to name novel products explicitly.

Our results align with, and extend, the high-income literature. The pooled estimate that social media tobacco exposure roughly doubles the odds of use (Donaldson et al., 2022) is mirrored by our adjusted estimate, which is striking given the contextual gulf between the United States and East Africa. The qualitative data add what meta-analyses cannot: a mechanism. Influencers function as credible peers, so their modelling of tobacco lowers perceived barriers and raises outcome expectancies — the observational-learning pathway long described in behavioural theory (Bandura, 1986) — and the absence of disclosure means followers process paid promotion as authentic endorsement, the very condition under which influence is strongest (Dhanesh and Duthler, 2019).

The protective factors are arguably the most policy-relevant results. Education, low tobacco-content exposure and self-reported resistance each independently reduced odds of use, and a majority of participants were already sceptical of influencer messaging and willing to be enlisted against it. This reframes urban youth not as passive targets but as latent allies — a more accurate and more hopeful premise for intervention than the deficit framing that dominates much prevention work.

### Strengths and limitations

The study’s strengths are its size (n=772), systematic sampling, comparative two-city design and methodological triangulation. Several limitations require candour. The cross-sectional design cannot establish temporality; we cannot exclude reverse causation, whereby tobacco users preferentially follow pro-tobacco influencers, and longitudinal work is needed to settle direction. Tobacco use was self-reported without biochemical verification (e.g. cotinine or exhaled carbon monoxide), risking social-desirability and misclassification bias, although anonymity and electronic self-completion likely reduced under-reporting. The content sample was purposive, hashtag-driven and limited to public posts, so it characterises rather than represents the universe of influencer content and probably understates exposure occurring in private or closed spaces. Finally, results from two highly connected capitals may not generalise to peri-urban or rural youth.

## Conclusion

Among urban youth in Kampala and Nairobi, exposure to influencers promoting tobacco was independently associated with tobacco use, operating alongside — not beneath — the established determinants of sex and age. The mechanism is not a hard sell but a soft one: tobacco woven into aspirational lifestyles by trusted peers, performing best when it is funniest and most emotive, and landing on an audience already primed to see vapes and shisha as the safer, smarter choice. The implication is direct. Tobacco control in East Africa cannot remain anchored to print-era advertising bans while the marketing has migrated to feeds those bans never imagined. Closing the gap means extending and enforcing prohibitions to cover influencer promotion, mandating disclosure of paid partnerships, holding platforms to proactive moderation of tobacco content, and — crucially — recruiting the same persuasive machinery for the other side, equipping credible young voices and media-literate audiences to make a smoke-free life the more aspirational story. The audience is already half-convinced; the task is to reach them in the language, and on the platforms, where the harm is currently being done.

## Acknowledgements

We thank the Centre for Tobacco Control in Africa for funding this research, and Makerere University College of Health Sciences, APAR Foundation, and the Students’ Campaign Against Drugs (Kenya) for institutional support. We are grateful to the research assistants and the 772 participants who shared their experiences, and to the Makerere University School of Public Health and KCA University Research Ethics Committees for their guidance.

## Data availability

De-identified data are available from the corresponding author on reasonable request, subject to ethical approval requirements.

## Author contributions

JSJK: conceptualisation, methodology, formal analysis, investigation, data curation, writing — original draft, project administration. TMRA, BO, KDL, SM: investigation, data collection, validation, writing — review and editing. JA and LN: supervision, methodology, writing — review and editing. All authors approved the final manuscript and meet ICMJE authorship criteria.

